# High-Dimensional Multinomial Multiclass Severity Scoring of COVID-19 Pneumonia Using CT Radiomics Features and Machine Learning Algorithms

**DOI:** 10.1101/2022.04.27.22274369

**Authors:** Isaac Shiri, Shayan Mostafaei, Atlas Haddadi Avval, Yazdan Salimi, Amirhossein Sanaat, Azadeh Akhavanallaf, Hossein Arabi, Arman Rahmim, Habib Zaidi

**Affiliations:** Division of Nuclear Medicine and Molecular Imaging, Geneva University Hospital, CH-1211 Geneva, Switzerland; Division of Clinical Geriatrics, Department of Neurobiology, Care Sciences and Society, Karolinska Institutet, Stockholm, Sweden; School of Medicine, Mashhad University of Medical Sciences, Mashhad, Iran; Departments of Radiology and Physics, University of British Columbia, Vancouver BC, Canada; Department of Integrative Oncology, BC Cancer Research Institute, Vancouver BC, Canada; Geneva University Neurocenter, Geneva University, Geneva, Switzerland; Department of Nuclear Medicine and Molecular Imaging, University of Groningen, University Medical Center Groningen, Groningen, Netherlands; Department of Nuclear Medicine, University of Southern Denmark, Odense, Denmark

**Keywords:** COVID-19, Pneumonia, Radiomics, Machine Learning, Computed Tomography, High-Dimensional Data

## Abstract

We aimed to construct a prediction model based on computed tomography (CT) radiomics features to classify COVID-19 patients into severe-, moderate-, mild-, and non-pneumonic. A total of 1110 patients were studied from a publicly available dataset with 4-class severity scoring performed by a radiologist (based on CT images and clinical features). CT scans were preprocessed with bin discretization and resized, followed by segmentation of the entire lung and extraction of radiomics features. We utilized two feature selection algorithms, namely Bagging Random Forest (BRF) and Multivariate Adaptive Regression Splines (MARS), each coupled to a classifier, namely multinomial logistic regression (MLR), to construct multiclass classification models. Subsequently, 10-fold cross-validation with bootstrapping (n=1000) was performed to validate the classification results. The performance of multi-class models was assessed using precision, recall, F1-score, and accuracy based on the 4×4 confusion matrices. In addition, the areas under the receiver operating characteristic (ROC) curve (AUCs) for multi-class classifications were calculated and compared for both models using “multiROC” and “pROC” R packages. Using BRF, 19 radiomics features were selected, 9 from first-order, 6 from GLCM, 1 from GLDM, 1 from shape, 1 from NGTDM, and 1 from GLSZM radiomics features. Ten features were selected using the MARS algorithm, namely 2 from first-order, 1 from GLDM, 2 from GLRLM, 2 from GLSZM, and 3 from GLCM features. The Mean Absolute Deviation and Median from first-order, Small Area Emphasis from GLSZM, and Correlation from GLCM features were selected by both BRF and MARS algorithms. Except for the Inverse Variance feature from GLCM, all selected features by BRF or MARS were significantly associated with four-class outcomes as assessed within MLR (All p-values<0.05). BRF+MLR and MARS+MLR resulted in pseudo-R^2^ prediction performances of 0.295 and 0.256, respectively. Meanwhile, there were no significant differences between the feature selection models when using a likelihood ratio test (p-value =0.319). Based on confusion matrices for BRF+MLR and MARS+MLR algorithms, the precision was 0.861 and 0.825, the recall was 0.844 and 0.793, whereas the accuracy was 0.933 and 0.922, respectively. AUCs (95% CI)) for multi-class classification were 0.823 (0.795-0.852) and 0.816 (0.788-0.844) for BRF+MLR and MARS+MLR algorithms, respectively. Our models based on the utilization of radiomics features, coupled with machine learning, were able to accurately classify patients according to the severity of pneumonia, thus highlighting the potential of this emerging paradigm in the prognostication and management of COVID-19 patients.

## INTRODUCTION

The highly contagious SARS-CoV-2 virus has led to significant morbidity and mortality worldwide ^1^. Pneumonia is regarded as one of the main complications of COVID-19 disease, which can lead to lethal conditions while escalating the cost of healthcare ^2^. The most popular diagnostic test considered as the gold standard for coronavirus disease is the reverse transcription polymerase chain reaction (RT-PCR) assay ^3^. While highly specific, RT-PCR has shown low sensitivity, as studies have reported significant false-negatives in patients who had abnormalities in their chest CT images confirmed with secondary follow-up RT-PCR to be positive for COVID-19 ^4^.

CT aids in the diagnosis and management of COVID-19 patients and could be potentially used as an outcome/survival prediction tool, towards enhanced treatment planning ^5-7^. CT scanning has been utilized as a highly sensitive tool for COVID-19 diagnosis ^8^ since it is fast and generates quantifiable features (e.g., the extent to which lung lobes are involved) and non-quantifiable features (e.g., ground-glass opacities and their laterality) to assess COVID-19 pneumonia, besides the enhanced sensitivity compared to RT-PCR ^9^.

Severity can be defined as an index that depicts the effects of a disease on mortality, morbidity, and comorbidities ^10^ and has the potential to help physicians manage the patients more decently whether in patients with cancer or with non-cancer diseases ^11,12^. A number of severity scoring systems have been proposed to quantify disease advancement in patients, including general assessments (e.g., APACHE score) and disease-specific ones (e.g., Child-Pugh score) ^13^. Several conventional scoring systems have been proposed for COVID-19 severity assessment ^14^. These include the usage of patient clinical, comorbidity, and laboratory data, which are all helpful in constructing predictive models for severity assessment in COVID-19 ^15^.

There has also been a growing interest in using imaging data of patients, such as thoracic CT images. For example, a study by Sanders et al. ^16^ computed the score of CT images in patients with cystic fibrosis and evaluated the prognostic ability. A promising line of research that emerged recently reported on the CT severity index and its correlation with acute pancreatitis severity ^17-19^. The COVID-19 Reporting and Data System (CO-RADS) was suggested for standardized visual assessment of COVID-19 pneumonia to enhance agreement between radiologists ^20^. This system includes features for the diagnosis of COVID-19 and consists of a 5-point scale for categorizing patient CT images. In addition, other guidelines aiming to reach consensus when interpreting COVID-19 suspected chest CT images were proposed ^21^. These guidelines are mostly based on visual assessment of images; e.g. the amount to which lung lobes are involved, the volume of which is infected, and anatomical assessments.

Francone et al. ^22^ reported a study on the correlation between CT score and the severity of coronavirus disease. Zhao et al. ^23^ also conducted research on the measurement of the extent to which lung lobes are infected and evaluation in COVID-19 patients’ prognosis. Li et al. ^24^ also confirmed the association between chest CT score and COVID-19 pneumonia severity. At the same time, most scoring systems involve visual assessment and hence are time-consuming ^23,24^. In this regard, medical image analysis using machine learning and radiomics has been applied to quantify features to tackle these main challenges ^25-35^.

The field of radiomics opens pathways for the study of normal tissues, cancer, and many other diseases, including potentially the newly emerging COVID-19 disease ^6,7,29,36-40^. Specifically, Xie et al. ^41^ evaluated the potential of a radiomics framework to diagnose COVID-19 from CT images. Di et al. ^42^ also studied whether radiomics features can help to distinguish between pneumonia of COVID-19 and that of other viral/bacterial causes. A number of studies reported on the application of radiomics analysis to CT images towards COVID-19 classification and prognostication^43^. Homayounieh et al. ^44^ assessed the prognostic power of CT-based radiomics features to determine severe and non-severe cases. In another study, Li et al. ^45^ proposed a radiomics model based on CT images and classified patients based on the criticality of their disease. A recent study by Yip et al. ^46^ applied a robust radiomics model to CT images to predict the severity of COVID-19 disease in patients. All above models pursued binary task performance, which reduced multiclass classification to two class approaches. However, in the real clinical triage situation, scoring systems consist of multi-class datasets. In the present study, involving a large cohort of patients, we aimed to construct a CT radiomics-based multi-class classification model to predict the severity of COVID-19 pneumonia.

## MATERIALS AND METHODS

### Data Description

Figure 1 presents the different steps performed in this study. All experiments were performed in accordance with relevant guidelines and regulations.

**Figure 1:**
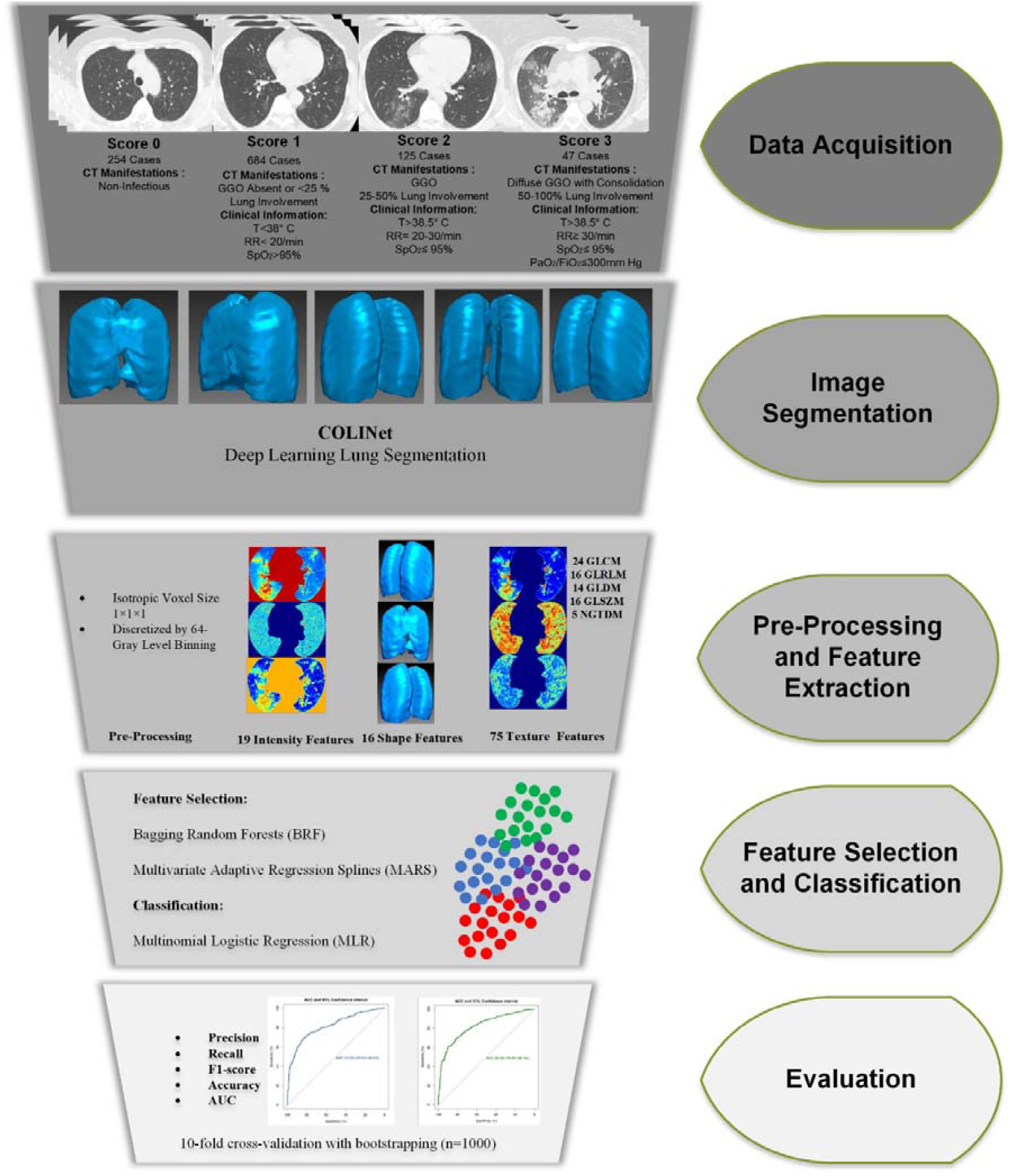
Different steps of current study. GGO : ground glass opacities, T: Temperature, RR: Respiratory Rate, SpO_2_: Peripheral Capillary Oxygen Saturation, PaO_2_: Partial Pressure of Oxygen. FiO_2_=Fraction of Inspired Oxygen.

### Datasets and Segmentation

This study is based on the MosMed Dataset (<u>mosmed.ai</u>) consisting of 1110 patient CT scans, also utilized in other efforts ^46,47^. Ethics approval and consent to participate **were** not needed since the study was preformed on open access online dataset. The patients were referred to the Municipal Hospital in Moscow, Russia, and were classified based on clinical and visual CT findings as follows.

In the zero class, the patient has neither clinical symptoms (e.g. fever) nor CT findings in favor of any kind of pneumonia (Class 0, non-pneumonic). The 1^st^ class contains patients who have a low-temperature fever (t < 38 °C) in addition to a mild increase in respiratory rate (RR <20) while showing none or < 25% ground-glass opacity (GGO) involvement (Class 1, COVID-19 with mild severity). Patients in the 2^nd^ class have a higher body temperature (t > 38.5 °C) with a RR of 20-30, while CT scan shows 25-50% involvement of lung parenchyma (Class 2, COVID-19 with moderate severity). Patients in the 3^rd^ class have high body temperature and RR of 30 or more, with CT findings of 50% to diffuse involvement in addition to organ failure and shock signs (Class 3, severe COVID-19). Each of the classes, namely 0, 1, 2, and 3, included 254, 684, 125, and 47 patients, respectively. The median age was 47 (ranging from 18 to 97), and 42% of patients were female. Figure 2 shows an example of representative CT images for each class.

**Figure 2:**
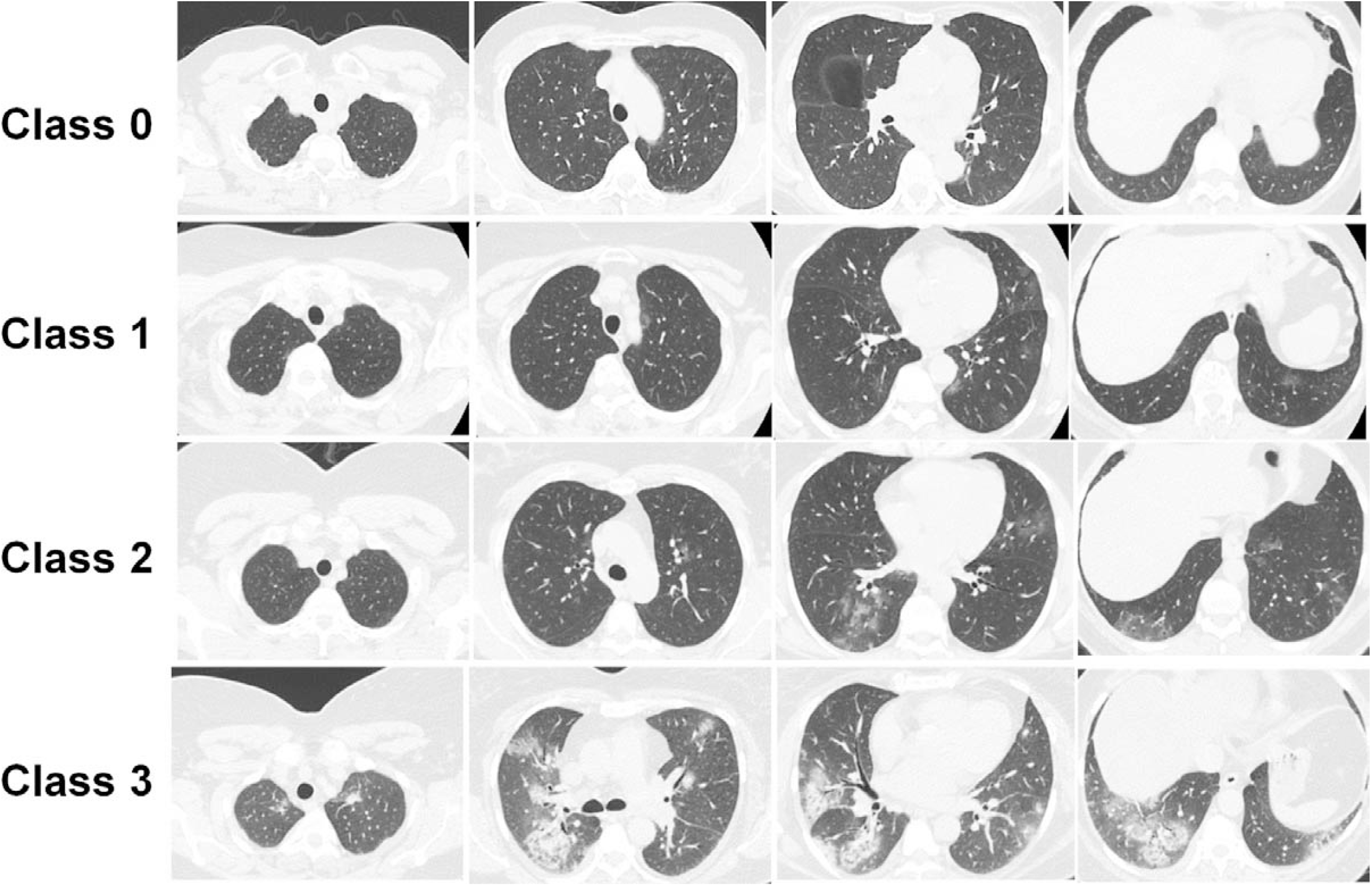
Example of patient CT images for different class.

All CT images were automatically segmented using a deep learning-based algorithm for whole lung segmentation ^48,49^. After whole-lung 3D segmentation, all images were reviewed and modified to ensure correct 3D-volume lung segmentation.

### Image Preprocessing and Feature Extraction

All images were resized to isotropic voxel size 1×1×1 mm^3^ and image intensity was discretized by 64-gray level binning, followed by feature extraction. The extracted features from the whole-lung segmented regions, totalling 110, included shape (n=16), intensity (n=19), and texture features, namely second-order texture of gray-level co-occurrence matrix (GLCM, n=24), and high-order features, namely gray-level size-zone matrix (GLSZM, n=16), neighbouring gray tone difference matrix (NGTDM, n=5), gray-level run-length matrix (GLRLM, n=16) and gray-level dependence matrix (GLDM, n=14). Radiomics feature extraction was performed using the Pyradiomics Python library ^50^, which is compliant with the Image Biomarker Standardization Initiative (IBSI) ^51^.

### Feature Selection and Classification and Evaluation

In this study, we used two different feature selection algorithms, including Bagging Random Forests (BRF) ^52^ and Multivariate Adaptive Regression Splines (MARS) ^53^. BRF and MARS algorithms were implemented in “VSURF” and “earth” R packages, respectively. For multiclass classification, we implemented multinomial logistic regression using the “mnlogit” R package. The MLR model fitness indices included p-value of the Wald test (corrected for false-discovery rate via Benjamini and Hochberg method), pseudo R^2^ (goodness of fit criteria in a logistic regression model), as well as coefficient and Standard of Error (SE). In the MLR model, class 0 served as a reference class whereas statistical comparison between two models (the two feature selectors) was performed by the Likelihood Ratio Test. Ten-fold cross-validation with bootstrapping (n=1000) was used to validate model performance. We report precision, recall, F1-score, and accuracy for different class for each model. In addition, the areas under the receiver operating characteristic (ROC) curve (AUCs) for multi-class classification models were calculated and compared for both models using “multiROC” and “pROC” R packages, respectively.

## RESULTS

Table 1 summarizes the selected features and their importance value (IV) by BRF and MARS for multiclass classification. Nineteen radiomics features were selected by BRF, including 9 from first-order, 6 from GLCM, one from GLDM, one from shape, one from NGTDM, and one from GLSZM. Among these features, Mean Absolute Deviation (IV: 80%), Robust Mean Absolute Deviation (IV: 72%) and kurtosis (IV: 70%) features from first-order, and Correlation (IV: 75%), and Cluster Tendency (IV: 73%) features from GLCM were selected as the most important ones. In the MARS algorithm, 10 features were selected with high IVs, including 2 from first-order, 1 from GLDM, 2 from GLRLM, 2 from GLSZM, and 3 from GLCM. The highest IV was achieved by Gray Level Variance from GLDM (IV: 94%), Zone Entropy from GLSZM (IV: 93%), and Small Area Emphasis from GLSZM (IV: 83%). Mean Absolute Deviation and Median from first-order features, Small Area Emphasis from GLSZM, and Correlation from GLCM, were selected by both BRF and MARS algorithms. Figure 3 depicts the feature map of different radiomics features in different classes. Figure 4 represents the feature selection process for multi-class classification by BRF and MARS.

**Table 1.** Selected features by Bagging Random Forests (BRF) (“VSURF” R package) and multivariate adaptive regression splines (MARS) (“earth” R package) for multi-class classification

| Feature Selection Algorithm | Selected Variables | Feature type | Importance Value |
| --- | --- | --- | --- |
| Bagging Random Forests | First Order | Mean Absolute Deviation | 80% |
|  | GLCM | Correlation | 75% |
|  | GLCM | Cluster Tendency | 73% |
|  | First Order | Robust Mean Absolute Deviation | 72% |
|  | First Order | Variance | 70% |
|  | First Order | Interquartile Range | 71% |
|  | First Order | Kurtosis | 72% |
|  | First Order | Skewness | 71% |
|  | GLDM | Dependence Entropy | 68% |
|  | First Order | Median | 48% |
|  | First Order | Entropy | 69% |
|  | GLCM | Sum Entropy | 67% |
|  | GLCM | Inverse Variance | 42% |
|  | GLCM | Joint Entropy | 66% |
|  | First Order | Uniformity | 63% |
|  | GLCM | Contrast | 62% |
|  | Shape | Major Axis Length | 30% |
|  | NGTDM | Contrast | 60% |
|  | GLSZM | Small Area Emphasis | 50% |
| Multivariate Adaptive Regression Splines | First Order | Mean Absolute Deviation | 22% |
|  | GLDM | Gray Level Variance | 94% |
|  | GLRLM | Gray Level Non Uniformity | 57% |
|  | GLRLM | Short Run Emphasis | 70% |
|  | GLSZM | Small Area Emphasis | 83% |
|  | First Order | Median | 24% |
|  | GLCM | Imc1 | 46% |
|  | GLSZM | Zone Entropy | 86% |
|  | GLCM | Correlation | 38% |
|  | GLCM | Difference Entropy | 40% |

**Figure 3:**
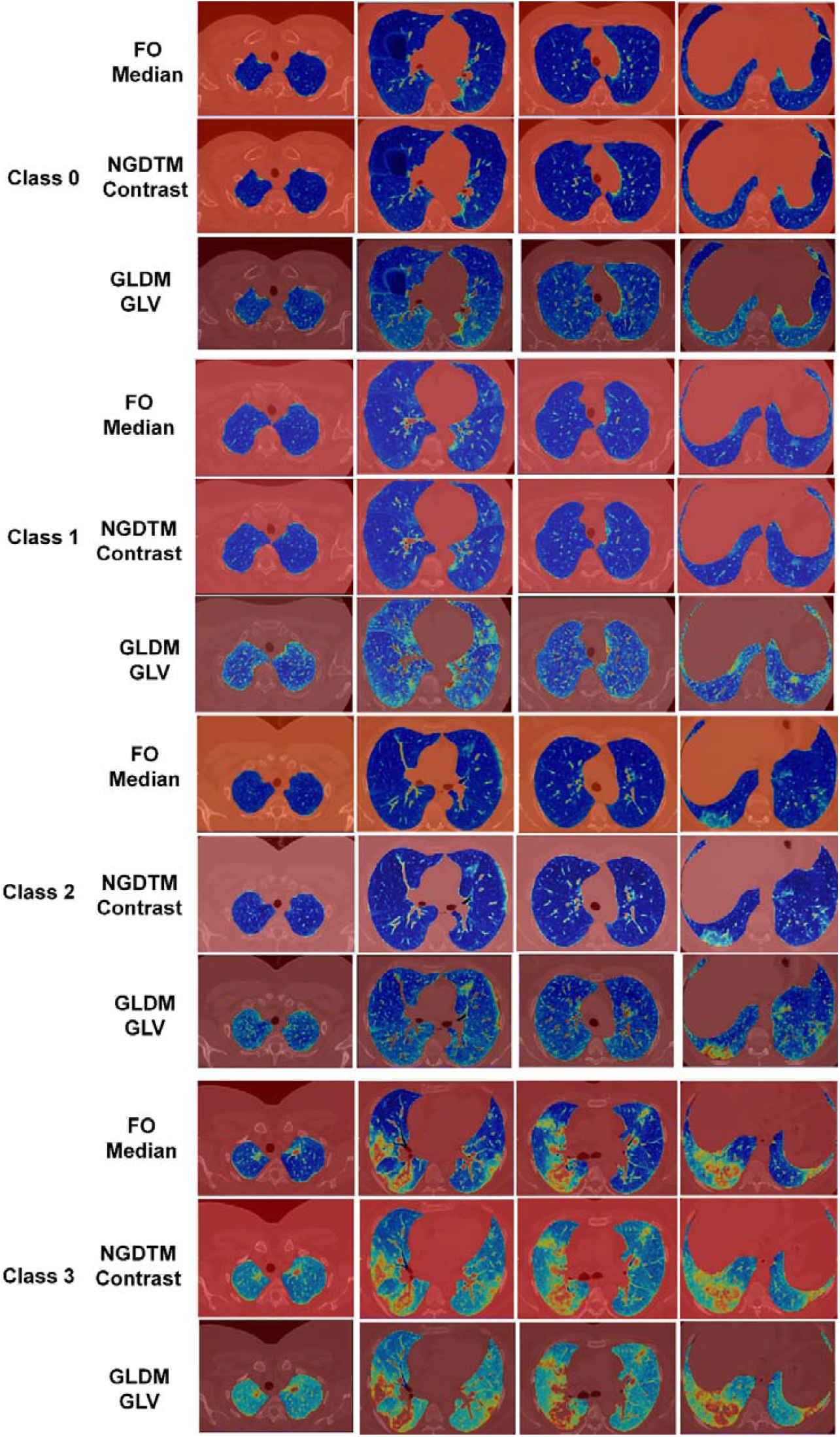
Example of selected features (Median feature from First Order, Contrast feature form NGDTM and GLV features from GLDM) in different class cases.

**Figure 4:**
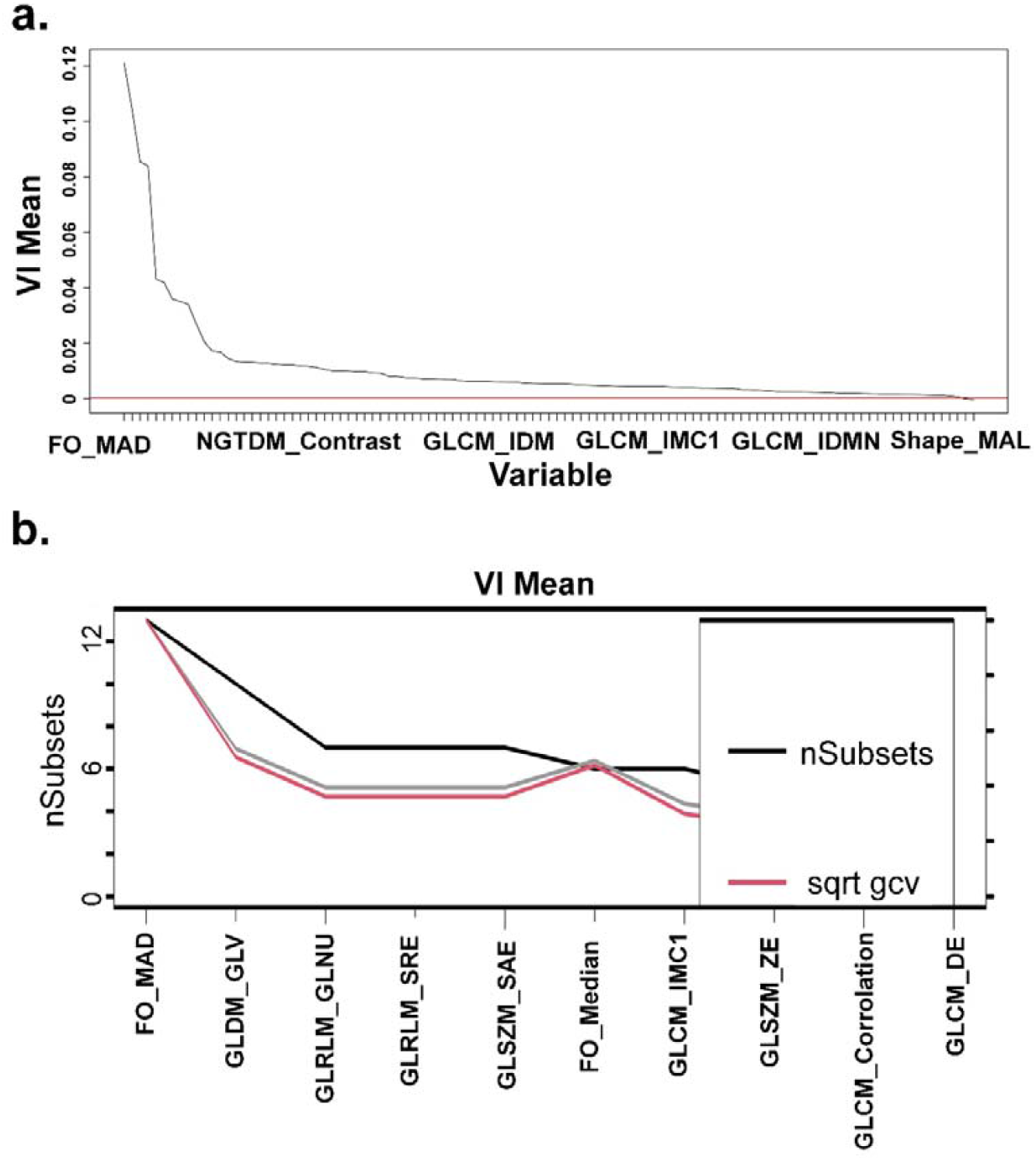
Feature selection process for multi-class classification by (a) Bagging Random Forests (number of selected features=19), and (b) Multivariate Adaptive Regression Splines (number of selected features=10).

Table 2 summarizes the adjusted p-value (by Benjamini and Hochberg method) of the Wald test and coefficient (standard of error) for selected features by BRF and MARS algorithms. Except for Inverse Variance from GLCM, all selected features yielded a significant p-value (<0.05). BRF+MLR and MARS+MLR resulted in pseudo R^2^ values of 0.295 and 0.256, respectively. However, there were no significant differences between both models when using a likelihood ratio test (p-value =0.319).

**Table 2.** Model fitness indices for application of multinomial logistic regression (MLR) (“mnlogit” R package) to selected features.

| Feature Selection Algorithm | Feature type |  | Adj. P-value | Coefficient (SE) | Pseudo R<sup>2</sup> for logistic regression |
| --- | --- | --- | --- | --- | --- |
| Bagging Random Forests | First order | Mean Absolute Deviation | 0.001 | 0.066 (0.007) | 0.295 |
|  | GLCM | Correlation | 0.001 | 14.3 (1.85) |  |
|  | GLCM | Cluster Tendency | 0.002 | 0.20 (0.024) |  |
|  | First Order | Robust Mean Absolute Deviation | 0.001 | 0.093 (0.10) |  |
| | First Order | Variance | 0.001 | $17 \times 10^{-4} (2 \times 10^{-5})$ | |
|  | First Order | Interquartile Range | 0.001 | 0.042 (0.0047) |  |
|  | First Order | Kurtosis | 0.002 | -0.35 (0.042) |  |
|  | First Order | Skewness | 0.001 | -2.44 (0.269) |  |
|  | GLDM | Dependence Entropy | 0.001 | 4.58 (0.585) |  |
|  | First order | Median | 0.002 | 0.022 (0.0031) |  |
|  | First order | Entropy | 0.001 | 4.43 (0.495) |  |
|  | GLCM | Sum Entropy | 0.001 | 4.14 (0.461) |  |
|  | GLCM | Inverse Variance | 0.790 | -2.17 (8.15) |  |
|  | GLCM | Joint Entropy | 0.001 | 2.42 (0.270) |  |
|  | First Order | Uniformity | 0.001 | -21.8 (2.60) |  |
|  | GLCM | Contrast | 0.002 | 1.18 (0.136) |  |
|  | Shape | Major Axis Length | 0.005 | -0.012 (0.016) |  |
|  | NGTDM | Contrast | 0.028 | 171.4 (20.60) |  |
|  | GLSZM | Small Area Emphasis | 0.031 | -50.2 (7.71) |  |
| Multivariate Adaptive Regression Splines | First Order | Mean Absolute Deviation | 0.001 | 0.066 (0.007) | 0.256 |
|  | GLDM | Gray Level Variance | <0.001 | 0.705 (0.082) |  |
| | GLRLM | Gray Level Non Uniformity | <0.001 | -0.006 ($8 \times 10^{-6}$) | |
|  | GLRLM | Short Run Emphasis | <0.001 | 42.2 (4.90) |  |
|  | GLSZM | Small Area Emphasis | 0.001 | -50.2 (7.0) |  |
|  | First Order | Median | 0.001 | 0.022 (0.003) |  |
|  | GLCM | Imc1 | 0.001 | -44.8 (6.82) |  |
|  | GLSZM | Zone Entropy | 0.001 | 6.3 (0.76) |  |
|  | GLCM | Correlation | 0.001 | 14.3 (1.85) |  |
|  | GLCM | Difference Entropy | 0.001 | 8.7 (0.98) |  |

P-value by Wald chi-square test; Adj. P-value: P-value adjusted by Benjamini and Hochberg false discovery (FDR) correction method; class 1 as a reference class; SE= Standard of Error; statistical comparison between two models showed non-significant difference by Likelihood Ratio Test: P-value=0.319.

Table 3 summarizes classification power indices (SD), including Precision, Recall, F1-score, Accuracy, and AUC via multinomial logistic regression with 1000 bootstrapping samples for each model. In terms of F1-score, classes 2 and 3 resulted in the lowest precision (mean (sd)) in BRF+MLR (0.798 (0.106)) and MARS+MLR (0.752 (0.099)), whereas four-class mean F1-scores were 0.847 and 0.805 for BRF+MLR and MARS+MLR algorithms, respectively. The mean precision was 0.861 and 0.825, whereas the mean recall was 0.844 and 0.793 for BRF+MLR and MARS+MLR algorithms, respectively. BRF+MLR and MARS+MLR algorithms achieved an accuracy of 0.933 and 0.922, respectively, in four-class classification. AUCs (95% CI) for multi-class classification were 0.823 (0.795-0.852) and 0.816 (0.788-0.844) for BRF+MLR and MARS+MLR algorithms, respectively. Figure 5 depicts the ROC curves for our four-class classification method.

**Table 3.** Classification performance indices by multinomial logistic regression with 1000 bootstrapping samples based feature selection. SD hown in brackets.

| Feature Selection Algorithm | Class | Precision | Recall | F1-score | Accuracy | AUC (95% CI) |
| --- | --- | --- | --- | --- | --- | --- |
| Bagging Random Forests | **Class 1** | 0.743<br>(0.078) | 0.948<br>(0.093) | 0.833<br>(0.099) | 0.912<br>(0.098) | 0.823 (0.795-0.852) |
|  | **Class 2** | 0.926<br>(0.022) | 0.861<br>(0.017) | 0.892<br>(0.017) | 0.872<br>(0.015) |  |
|  | **Class 3** | 0.866<br>(0.101) | 0.739<br>(0.099) | 0.798<br>(0.106) | 0.958<br>(0.110) |  |
|  | **Class 4** | 0.906<br>(0.10) | 0.829<br>(0.097) | 0.866<br>(0.096) | 0.989<br>(0.104) |  |
|  | **Average/total** | 0.861 | 0.844 | 0.847 | 0.933 |  |
| Multivariate Adaptive Regression Splines | **Class 1** | 0.732<br>(0.095) | 0.862<br>(0.101) | 0.792<br>(0.081) | 0.895<br>(0.126) | 0.816 (0.788-0.844) |
|  | **Class 2** | 0.901<br>(0.019) | 0.865<br>(0.014) | 0.883<br>(0.026) | 0.859<br>(0.017) |  |
|  | **Class 3** | 0.824<br>(0.079) | 0.764<br>(0.111) | 0.793<br>(0.091) | 0.955<br>(0.123) |  |
|  | **Class 4** | 0.842<br>(0.112) | 0.680<br>(0.108) | 0.752<br>(0.099) | 0.980<br>(0.106) |  |
|  | **Average/total** | 0.825 | 0.793 | 0.805 | 0.922 |  |

**Figure 5:**
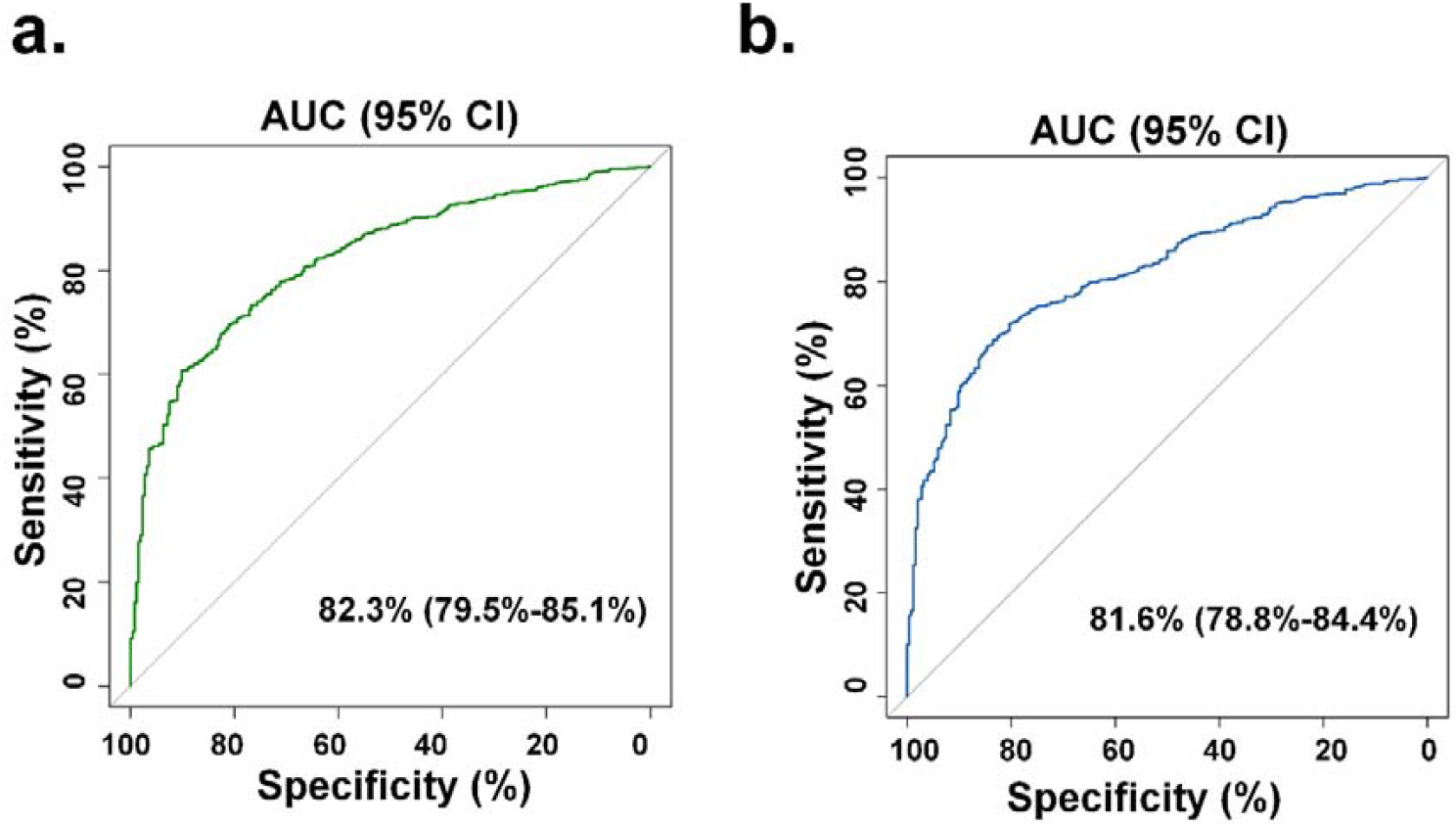
(a) ROC curve for assessing power of multi-class classification of the selected features in Bagging Random Forests (AUC=0.823), and (b) Multivariate Adaptive Regression Splines (AUC=0.816). Statistical comparison of ROC curves by “pROC” R package indicated non-significant difference (Z=-1.164, P-value=0.244).

## DISCUSSION

In the current study, we constructed a CT radiomics-based model to predict the severity of COVID-19 patients in a large cohort of patients. To this end, we extracted radiomics features from whole lung segmentations and selected high-importance features utilizing two different algorithms, namely BRF and MARS. The selected features were then fed to a multinomial logistic regression classifier for multiclass severity scoring. We achieved 0.823 (95% CI: 0.795-0.852) and 0.816 (95% CI: 0.788-0.844) for AUC, and 0.933 and 0.922 for accuracy in BRF- and MARS-selected features, respectively.

We used an automatic model ^48^ to segment chest CT images for two reasons. First, most CT scans performed in the COVID-19 pandemic era are low-dose. In addition, these scans are acquired with a high pitch. Hence, it is difficult for radiologists to find and follow lung fissures to manually detect or segment the anatomical lobes. As such, we used our previously constructed deep learning model to fully segment the entire lung of each patient.

Yip et al. ^46^ conducted a study on the same dataset utilized in this work, aiming to evaluate some radiomics features towards severity class prediction in patients. They included all 1110 patient CT scans and extracted 107 radiomics features. The maximum relevance minimum redundancy (MRMR) and recursive feature elimination (RFE) algorithms were exploited for feature selection and analysis of the selected features using univariate and multivariate approaches using a logistic regression model to classify as accurately as possible. In their study, the patients were categorized into three severity categories, namely mild, moderate, and severe, to perform two-class classification tasks (mild vs. severe and moderate vs. severe) by splitting the data into training (60%) and test (40%) sets. The authors obtained an AUC of 0.65 in differentiating between moderate and severe cases, while their model performed better (AUC = 0.85) in distinguishing mild vs. severe forms of COVID-19 disease. In this work, we reached an overall AUC of 0.823. In our study and the one by Yip et al. ^46^, feature extractions were performed using Pyradiomics ^50^ as applied to the entire lung. Interestingly, there were some commonly selected features arrived at via feature selection in both studies, including Small Area Emphasis from GLSZM, Correlation and Informational Measure of Correlation from GLCM, and Median and Mean Absolute Deviation from first-order features. These selected features in both studies could potentially be used as predictors as they provide information about the intensity and heterogeneity of the lung in COVID-19 patients.

A noticeable advantage of the study by Yip *et al*. ^46^ was the use of a second radiologist observer who classified patients’ images into mild, moderate, and severe classes without paying attention to the default classification of the dataset provider. This method helped to observe the prediction power of the models in both “provider” and “radiologist” datasets. In addition, they split the dataset into training and test sets. In contrast, we applied the bootstrapping technique to estimate and ensure the reproducibility of our results. In addition, the study by Yip *et al*. ^46^ may have reduced generalizability as it only predicts mild versus severe, and moderate versus severe disease, having reduced multiclass classification into two-class approaches. In the real clinical triage situation, the radiologist may benefit from a multiclass classification scheme for enhanced patient management, as provided by our study.

Homayounieh *et al*. ^54^ included 315 patients in their study and extracted CT-based radiomics features from the lung to show that radiomics can predict patients’ outcome (inpatient vs. outpatient management) with an AUC of 0.84 while the radiologist assessment alone achieved an AUC of 0.69. Feature extraction was performed by applying the different preprocessing algorithms on images, with classification performed using logistic regression. They reported that adding clinical variables to the radiomics model can notably improve the predictability of a model for patient outcome prediction (AUC improved from 0.75 to 0.84). Another study conducted by Wei *et al*. ^55^ evaluated the predictive ability of two models (one CT texture-based and one clinical) for determining the severity of each of the 81 COVID-19 patients. They showed that CT texture features could modestly predict whether the patient has common COVID-19 pneumonia or a severe one with an AUC of 0.93, which is comparable to that of the clinical-only model (AUC = 0.95). They also observed that several texture features had a moderate correlation with the clinical variables of patients.

Chaganti *et al*. ^56^ studied Ground Glass Opacity (GGO) and consolidations that appear on a CT image of COVID-19 patients in an attempt to propose an automated method for segmenting and quantifying COVID-19 lesions. Their proposed method calculated the percentage of opacity and lung severity score using deep learning algorithms and was able to predict the severity with a decent performance. However, Chaganti *et al*. ^56^ proposed a method trained only on the mentioned abnormalities and had a limited performance in other abnormalities quantification. Even with improving segmentation algorithms, this method would be limited because of the highly heterogeneous nature of COVID-19 pneumonia in addition to ignoring the shape and texture of segmented lesions. Moreover, providing accurate lobe segmentation of COVID-19 patients would be challenging from typical low-dose and high pitch chest CT scans. In the current and previous studies ^44,46,55^, radiomics features, as extracted from the entire lung (less challenging segmentation task for deep learning algorithms), were evaluated to provide fast and robust severity scoring in COVID-19 patients.

In this work, chest CT was used for assessment. At the same time, there are few studies on other modalities such as chest X-ray radiography in prognostication and outcome prediction evaluation of COVID-19 patients. For example, Bae and colleagues ^57^ utilized radiomics features and modeled them on chest X-rays of 514 patients and found out that their radiomics- and deep learning-based model can accurately predict mortality and the need for mechanical ventilation in patients (AUCs = 0.93 and 0.90, respectively). Providing a severity score using chest X-rays is a valuable venue to explore. Yet, such work requires extensive comparisons with CT-based frameworks to assess the relative value of each modality for different tasks.

This study suffered from a few limitations, including the fact that our model was trained on single-center data. At the same time, we evaluated our models using a 10-fold cross-validation and bootstrapping technique to evaluate the repeatability and robustness of our results. In any case, further research should be conducted on multicentric data and patient images with multiple observers for improved training of the models and enhanced generalizability.

## Conclusion

We evaluated high-dimensional multinomial multiclass severity scoring of pneumonia using CT radiomics features and machine learning algorithms. We applied two feature selectors (BRF and MARS) coupled to one classifier (multiclass logistic regression model) on a large cohort of COVID-19 patients. Our radiomics model was validated to depict accurate classification of patients according to multi-class pneumonia severity assessment criteria, highlighting the potential of this emerging paradigm in the assessment and management of COVID-19 patients.

## Data Availability

All data produced are available online at

https://mosmed.ai/datasets/covid19_1110

## Acknowledgments

This work was supported by the Swiss National Science Foundation under grant SNRF 320030_176052.

## Abbreviations

CT: Computed Tomography
COVID-19: Coronavirus disease 2019
AUC: Area under the receiver operating characteristic curve
BRF: Bagging Random Forest
FS: Feature Selection
GGO: Ground Glass Opacity
IBSI: The Image Biomarker Standardization Initiative
MARS: Multivariate Adaptive Regression Splines
MLR: Multinomial Logistic Regression
RT-PCR: Reverse transcription polymerase chain reaction
GLCM: Gray-Level Co-Occurrence Matrix
GLSZM: Gray-Level Size-Zone Matrix
NGTDM: Neighbouring Gray Tone Difference Matrix
GLRLM: Gray-Level Run-Length Matrix
GLDM: Gray-Level Dependence Matrix

